# Quantifying the Dynamics of Migration after a Disaster: Impact of Hurricane Maria in Puerto Rico

**DOI:** 10.1101/2020.01.28.20019315

**Authors:** Rolando J. Acosta, Nishant Kishore, Rafael A. Irizarry, Caroline Buckee

**Affiliations:** Department of Biostatistics, Harvard University, Cambridge, MA, USA; Center for Communicable Disease Dynamics, Department of Epidemiology, Harvard T.H. Chan School of Public Health, Boston, Massachusetts, United States of America; Department of Data Sciences, Dana-Farber Cancer Institute, Boston, MA, USA

**Author notes:** These individuals contributed equally to this work.

## Abstract

Population displacement may occur after natural disasters, permanently altering the demographic composition of the affected regions. Measuring this displacement is vital for both optimal post-disaster resource allocation and calculation of measures of public health interest such as mortality estimates. Here, we analyzed data generated by mobile phones and social media to estimate the weekly island-wide population at risk and within-island geographic heterogeneity of migration in Puerto Rico after Hurricane Maria. We compared these two data sources to population estimates derived from air travel records and census data. We observed a loss of population across all data sources throughout the study period, however, the magnitude and dynamics differ by data source. Census data predict a population loss of just over 129,000 from July 17 to July 2018, a 4% decrease; air travel data predicts a population loss of 168,295 for the same period of time, a 5% decrease; mobile phone based estimates predicts a loss of 235,375 form July 2017 to May 2018, an 8% decrease; and social media based estimates predict a loss of 476,779 from August 2017 to August 2018; a 17% decrease. On average, municipalities with smaller population size lost a bigger proportion of their population. Moreover, we infer that these municipalities experienced greater infrastructure damage as measured by the proportion of unknown locations stemming from these regions. Finally, our analysis measures a general shift of population from rural to urban centers within the island.

## Introduction

In the aftermath of natural disasters both short-term population displacement and longer-term migration may occur, leaving some affected regions permanently altered (1–3). Measuring population displacement is a priority in the immediate days and weeks after an event like a hurricane or flood, for the provision of aid and other supplies to communities in need. It is also critical to inform mortality estimates and other measures of public health interest that require an up-to-date denominator, since census estimates may be rendered inaccurate (4, 5). Measuring shifts in demographic composition and the geographic distribution of populations on longer timescales is also critical to rebuilding efforts, and to the development of frameworks for building resilience to future disasters. Currently, however, there are few data sources that can be used to rapidly assess and monitor population displacement in the short- and medium-term timescales after disasters happen.

In the absence of reliable migration data in the wake of natural disasters, the population size estimate used by government agencies and researchers generally rely on census estimates, and assume a linear change in population size between intervals or a constant population size since the most recent estimate (6). New approaches to estimating fluctuating denominators in near real-time would greatly improve disaster response and the assessment of local needs in the short- and long-term aftermath. Rapid censuses conducted in short intervals before and after a disaster are both logistically and financially impractical. In an increasingly digitally connected world, however, passively collected digital records are often maintained by technological services providers for billing or marketing purposes. These data, such as flight information, mobile phone data, or social media traces, can provide insight into the fluctuation of their respective populations at a high temporal and geographic resolution, both before and after a disaster. Assuming that appropriate steps are taken to anonymize and aggregate these data streams in secure ways, novel data streams offer ways to assess the needs of populations more accurately (7, 8).

Hurricane Maria made landfall in Puerto Rico as a category 4 storm on September 20th, 2017, becoming the third costliest hurricane in United States history (9, 10). In the ensuing weeks, the damage to infrastructure caused by the storm resulted in widespread lack of access to electricity, communication and health services (11, 12). Population displacement off the island and within Puerto Rico was widespread, although this was difficult to monitor directly. Direct and indirect mortality caused by the storm also increased in the months after the hurricane (11–16), but estimating mortality was made more complicated by the migration of populations because the population-at-risk in different parts of the island were shifting over time. Due to Hurricane Maria, the U.S Census Bureau ceased operations of the Puerto Rico Community Survey (PRCS) (a monthly survey of 36,000 housing units across the island) from October through December of 2017. Operations were resumed in January of 2018 with early results showing an island-wide increase in out-migration in 2017 compared to 2016 (17). The most recent updates from the PRCS show a continued increase in out-migration with a general population that decreased by 4% from 2017 to 2018 (18).

In this study, we evaluate two passively collected data sources and compare them to data on air travel and census data, to evaluate the effects of Hurricane Maria on large scale population fluctuation in Puerto Rico. We investigate the ability of these data sources to estimate island-wide population at risk post disaster and identify within-island geographic heterogeneity in population migration after the storm. We observe a non-linear change in population at-risk following Maria, and show that this difference is affected by rurality. These passively collected data sources also provide insight into regions that are more heavily affected by disasters and can augment the resources available to first responders. Each data source has its own limitations and biases, and could be used in conjunction with traditional census-based population estimates to improve the response to natural disasters, and to understand how to build more resilient systems in anticipation of future events.

## Results

### Data Sources

We compared four independent datasets to estimate population changes over the course of a year after the hurricane. First, we obtained the intercensal yearly population estimates provided by the American Community Survey (ACS) for 2010 to 2018, a yearly survey conducted by the US Census Bureau. The ACS estimates are for July 1st of each year.

Second, we extracted data from *Disaster Maps*, provided by Facebook’s Data for Good team. Specifically, for a group of people determined to be living in Puerto Rico the week before Hurricane María, weekly estimates from August 21st, 2017 to July 30th, 2018 on the number of these users residing in 77 out of the 78 municipalities on the island were available. Note that this data is a closed cohort of known individuals, so no new users appear in the data and there is a constant rate of attrition of users expected. However, this is also the only data set on a municipality, rather than an island-wide, level.

Third, Teralytics provided island-wide daily population proportion estimates from May 31, 2017 to April 30, 2018 based on cell phone usage patterns, analyzed in partnership with an unnamed mobile operator on the island. Since we don’t know which operator the data are from, it was impossible to assess the geographic bias in ownership in this group. The proportions were calculated relative to a baseline population determined by Teralytics using a method unavailable to the authors. Due to the unreliability of data for the four weeks after hurricane Maria, presumably due primarily to low connectivity, proportion estimates were not provided for this time period.

Finally, we obtained Airline Passenger Traffic data from the US Bureau of Transportation Statistics (BTS) through the Puerto Rico Institute of Statistics. The data is composed of monthly counts of passengers that arrived and left the island per month from January 2010 to February 2018. This data is unbiased but has a coarse temporal and geographic scale, and cannot account for the same individuals on repeated trips or distinguish Puerto Rican residents from short-term aid workers and other visitors.

Details on how each of these datasets were constructed are included in the ***Methods*** section and a side-by-side comparison is found in Supplemental Table 1.

**Table 1:** Comparison of Data Sources

| **Table 1: Comparison of Data Sources** |  |  |  |  |
| --- | --- | --- | --- | --- |
| **Data Source** | **Access** | **Time to Availability Post-Disaster** | **Cohort** | **Counting Algorithm** |
| Aviation Records | Monthly report by the U.S. Bureau of Transport Statistics published online. | Months | Open cohort of all individuals who took flights in and out of Puerto Rico. | Register of all flights in and out of Puerto Rico with lists of total number of passengers. |
| Census (ACS) | ACS yearly update between census years provided by the U.S. Census Bureau. | Depends on proximity to release date. Released yearly. | Open cohort of all individuals who reside in sampled households. | Monthly survey of a sample of households used to provide estimates of larger geographic regions. |
| Facebook | Published online through a Facebook portal as requested by researchers. | Near real time and generally open to most researchers. | Closed cohort of people who used Facebook located in Puerto Rico during the five weeks before Hurricane Maria and who logged in through a browser allowing for capture of their IP. | Cities are defined internally and a home city is assigned to each individual. Computation is performed in terms of transition counts from recorded "home_city" to "current_city" pairs. Only transition counts that are greater than 100 are retained. Transition matrices are summed to provide total number of Facebook users present in a city, number of users for whom that city is their "home_city", number of users present in the city for whom it is not their "home_city" and number of users whose location is unknown. |
| Call Detail Records | Provided by Teralytics in response to data request as a partnership. | ~1 week following aggregation and internal calculations. | Open cohort of cell phone users from a major undisclosed telecommunications operator in Puerto Rico. | Use of a proprietary model to extrapolate population estimates. |

### Population decreased after Hurricane María

We found agreement across all data sources of a consistent loss of population from July 1, 2017 to July 1, 2018, however, the dynamics and magnitude of the loss differs between data types (Figure 1). The ACS predicts a population loss of 129,848, a 4% decrease; the Airline Passenger Traffic data predicts a population loss of 168,295, a 5% decrease; the Teralytics data predicts a population loss of 235,375, an 8% decrease; and the Facebook data predicts a 17% decrease, which equates to a total estimated population loss of 475,779 on the island. In all cases, we observed a sharp drop after the hurricane. Both the Airline Passenger Traffic data and Teralytics data show a rebound in population, and stabilization, after December 31, 2017. The Facebook data does not show stabilization until April 2018. We note that the Facebook data is based on a closed cohort that is not able to measure population increases due to immigration to Puerto Rico. However, it does represent the cohort that likely experienced Hurricane Maria.

**Figure 1:**
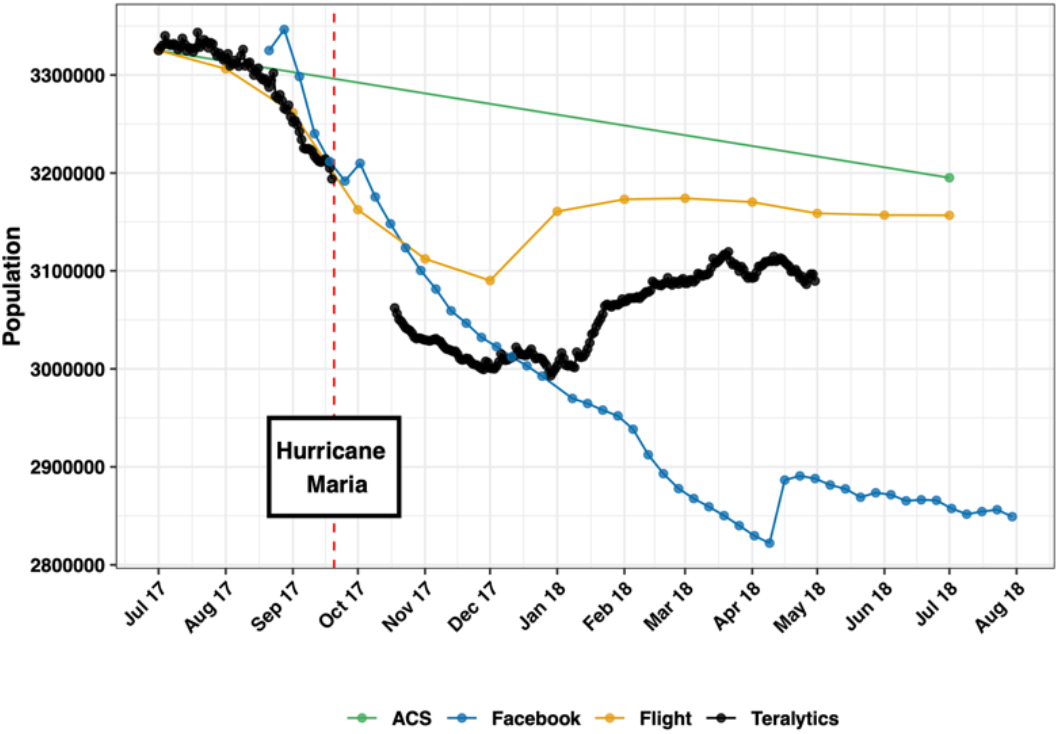
Population estimates for each data source.

### Small towns lost a larger portion of their population compared to large ones

Facebook provided municipality level data that allowed us to assess within island migration. For each municipality, the data included weekly counts of individuals still in the municipality and new to the municipality. For privacy protection, the new-to-municipality data was not included (see the ***Methods*** section for details on the data imputation techniques used, as well as a sensitivity analysis comparing different imputation approaches). On average, municipalities with smaller population size loss a bigger proportion of their population during the study period (Figure 2). The municipality of Toa Baja was a large outlier (Supplemental Figure 1) due to a reported surge of new individuals moving there at the end of the study period. Possible explanations for Toa Baja being an outlier are provided in the ***Discussion*** section. Apart from this outlier, San Juan, which is the capital of Puerto Rico, is the only municipality with more individuals at the end of the study period relative to baseline.

**Figure 2:**
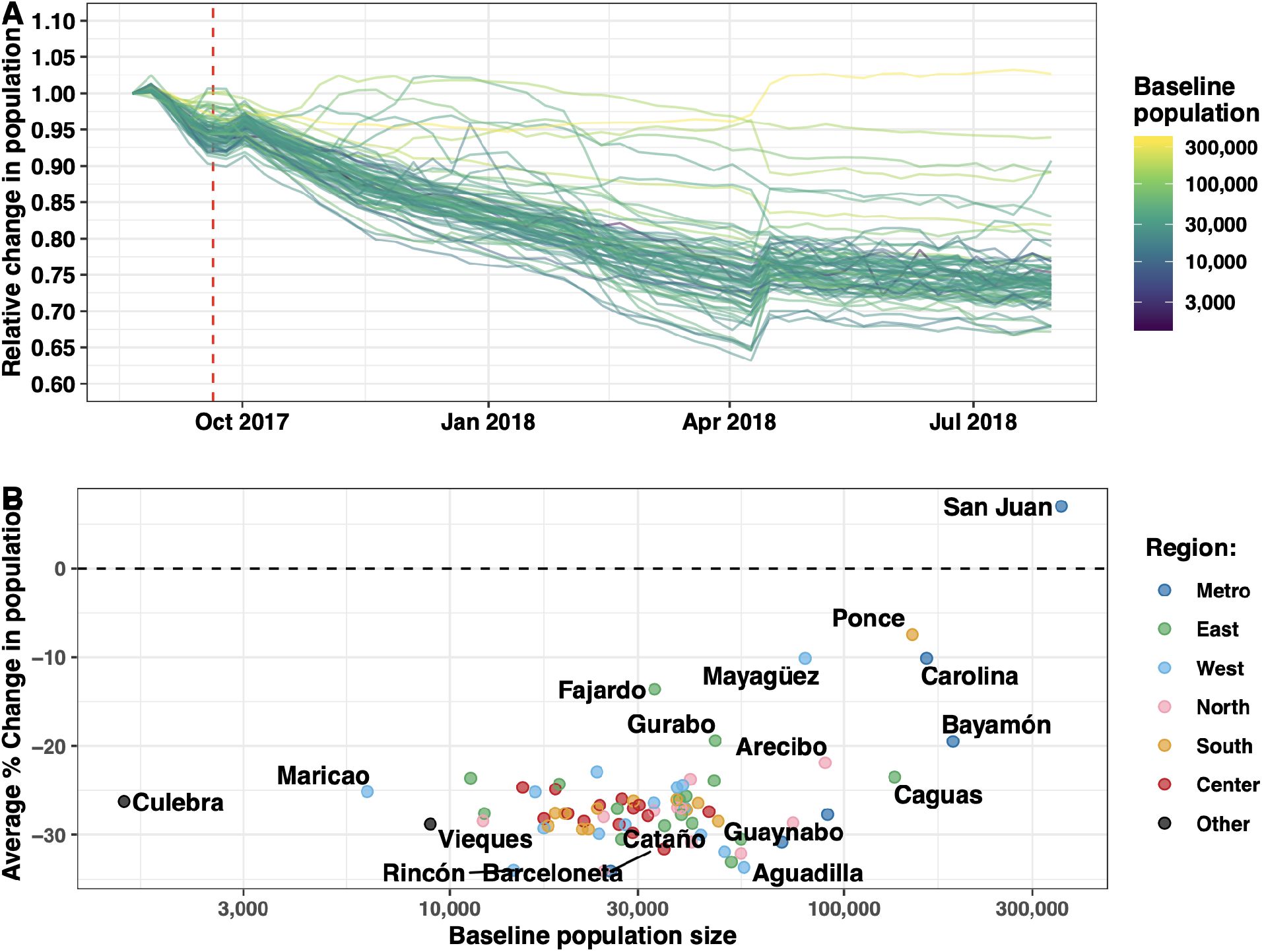
Small towns lost a larger portion of their population compared to large ones. (A) Relative change in the population of Facebook cohort member per municipality. Each curve corresponds to a different municipality, Population size in July 2017 is denoted by color. (B) Average percent change in population at the end of the study period, relative to baseline, compared to the ACS population size of municipality at baseline. The geographical regions are presented in Supplemental Figure 4. We fitted a linear model to each population curve and computed the average percent change using the first and last fitted values. The vertical lines correspond to the day when Hurricane Maria made landfall in Puerto Rico.

### Within island migration shifted populations from rural to urban regions

All municipalities experienced loss of their baseline resident populations. Much of this loss is explained by off-island immigration (Figure 3). However, the locality gaining the most new residents was San Juan (Figure 3). In fact, for urban areas, the baseline resident population loss was compensated by in-migration from other municipalities. For example, San Juan experienced a 19% loss of it’s baseline resident population but ended the study period with a 7% increase in total population suggesting in-migration of 26% by the end of the study period. These results taken together suggest a migration pipeline from rural to urban municipalities and likely off-island.

**Figure 3:**
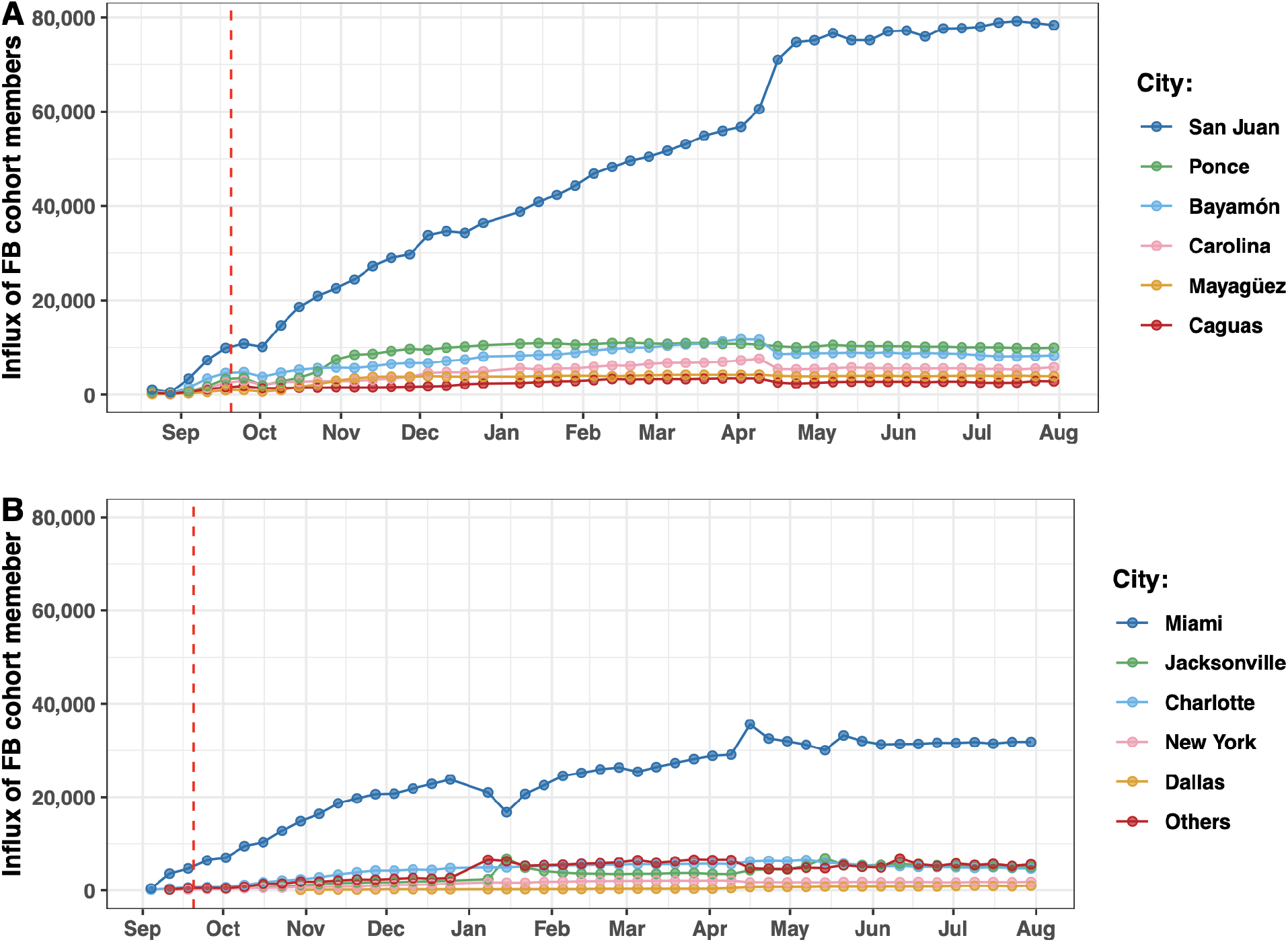
Immigration of Puerto Rican FB cohort members within and outside of Puerto Rico (A) Top 6 municipalities in Puerto Rico in terms of influx of Facebook cohort members who were located elsewhere before Hurricane Maria. The “Others” curve corresponds to the influx of all other municipalities of the island together. (B) Top 5 destinations of Facebook cohort members after Hurricane Maria. The “Others” curve correspond to the influx of all other destinations together. The vertical lines correspond to the day when Hurricane Maria made landfall in Puerto Rico.

### Infrastructure damage was greater in rural areas

Disaster Maps provided information about the proportion of people whose location is unknown for a particular week. Unknown locations may be the result of a) people stopping use of Facebook, b) people have changed their Facebook behavior, or c) loss of electricity and communication infrastructure. In the first full week of data collection immediately after the hurricane, approximately half of the cohort members did not register a location (Figure 4a), likely due to the widespread loss of infrastructure. During this week, the proportion of users with reported unknown location was substantially higher in the rural areas compared than in urban areas (Figure 4b, Supplemental Figure 2).

**Figure 4.**
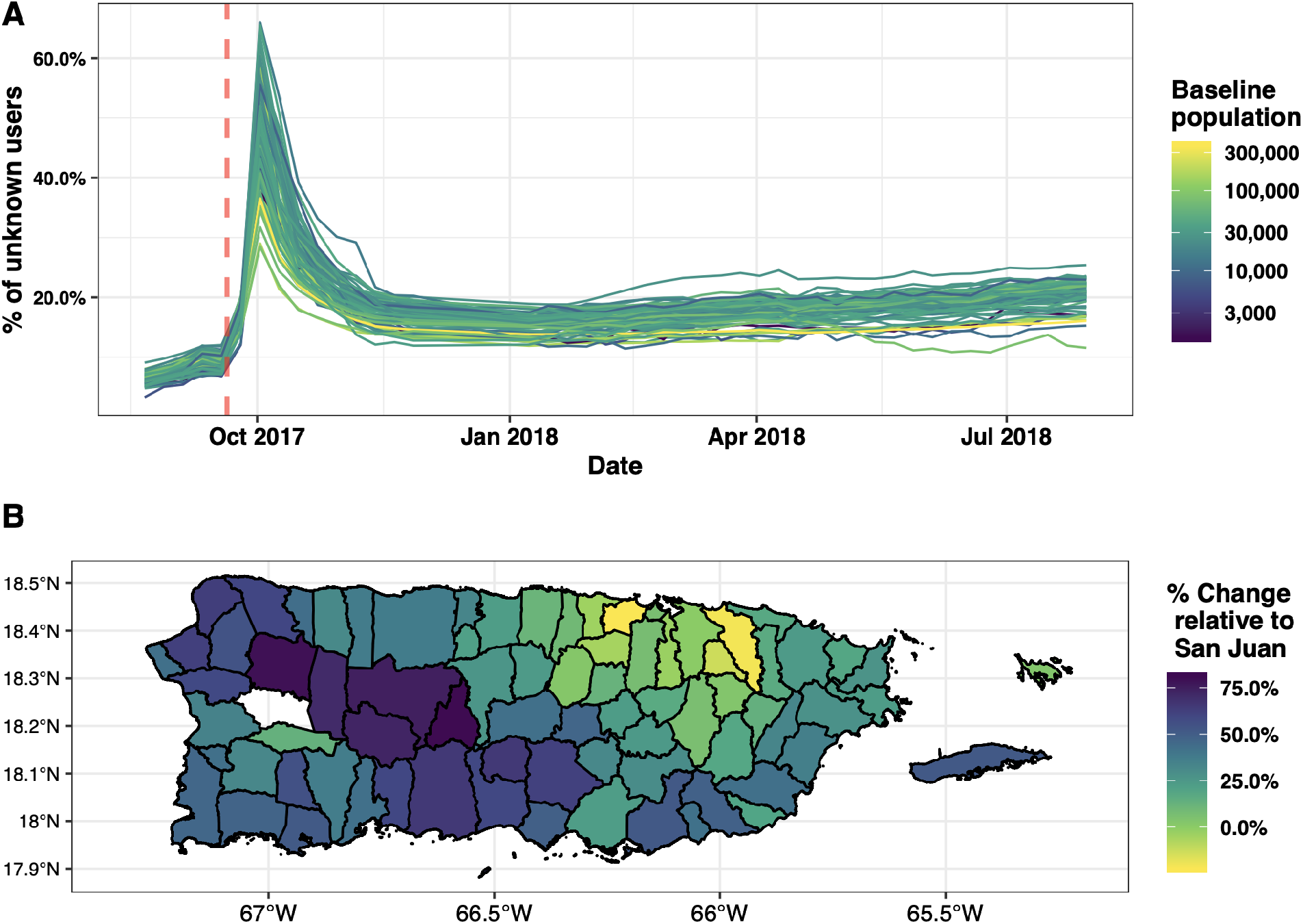
Facebook users with unknown locations. (A) Percent of individuals whose location was unknown by municipality. The vertical line denotes the day Hurricane Maria made landfall. Baseline population size is denoted in color.(B) Percent of individuals with unknown location relative to San Juan for the week of October 2nd, 2017, the week after Hurricane Maria. Note that we have no data for Las Marias.

In Supplemental Table 2, we show the top 15 municipalities which had a greater proportion of cohort members with their location unknown compared to the island-wide average. While rurality is directly linked with availability of infrastructure, the Facebook data highlight the heterogeneity in loss of access to resources in the immediate aftermath of a disaster. Some municipalities continue to have missing data relative to the island-wide average for many months

## Discussion

We show that new data sources for estimating population displacement may provide insights on the dynamics of migration following a natural disaster that cannot be obtained using traditional methods. Our results point to a consistent and long term loss of population in Puerto Rico after hurricane Maria as well as a rural-urban shift in populations that stayed on the island. In both the Facebook and Teralytics data we see decreases in population that start a few weeks prior to Hurricane Maria, likely due to the passing of Hurricane Irma a week before. In the Teralytics data and the flight data, the decrease in population levels off in December 2017 and in January 2018 we see a rebound in Puerto Rico’s population. In the Facebook data, the population loss continues until April 2018, resulting in the lowest overall population estimate. Both datasets provided island-wide population estimates that were lower than the ACS vintage 2018 point estimate. Although each has its own biases, these passively-collected data estimates follow similar trends and show dynamic population fluctuations.

Using the Facebook data to analyze within-island movements, we observed persistent migration from rural to urban areas of Puerto Rico. This may be explained by individuals migrating from rural to urban areas in search of basic needs in the short term and staying due to increased access to resources in the long term. Out-migration from rural areas continued in the Facebook cohort over the course of the available data, becoming more concentrated in urban areas. Previous household surveys suggest greater out-migration among younger individuals, potentially changing the demographic distribution of rural versus urban regions in important ways (12).

In the immediate aftermath of a disaster, electricity, communication and transportation are often affected leading to sparse information on the areas most heavily affected. Assuming that individuals will continue to use services like mobile phones and Facebook if they had access, the lack of interaction with these services in the immediate aftermath of these services could serve as a proxy for damage from the hurricane. Spatial and temporal granularity in these immediately available datasets could augment satellite imagery and primary data sources to more readily target priority areas for response. In the Facebook data, we can see that in the weeks immediately following Hurricane Maria, we identify areas which had larger proportions of cohort members whose locations were unknown compared to San Juan. These areas coincide with rural municipalities found to be some of the first declared as disaster zones post hurricane (19).

Passively-collected data provide a promising supplement to current at-risk population estimation procedures, however each data source has its own biases and limitations. The population estimates from Facebook versus Teralytics diverged significantly both before (by >9,500 people on September 20, 2017) and after (by >210,000 people on April 20, 2018) the hurricane. These data are not necessarily comparable, since neither the market share of mobile phone providers nor Facebook users are likely to be perfectly representative of the overall population. In general, we expect those not included in the Facebook sample to be those without phones or internet access, which would likely include children, the elderly, and the very poor (20). For privacy reasons, we were unable to analyze the demographic composition of the Facebook cohort, but these data represent a meaningful proportion of the Facebook population (21). The Facebook data also represents a closed cohort, unlike the other sources. Membership into this group is defined during the 5-week, pre-crisis period and no individuals are able to enter afterwards. This means that this cohort will inherently only decrease in size. This rate of decrease is driven by 1) The baseline mortality rate, 2) the excess mortality due to the crisis, and; 3) the baseline rate of discontinuation of Facebook use. One important advantage of the Facebook data is the ability to analyze within-island movement patterns, which is not possible using flight or Teralytics estimates. Teralytics analyzed mobile phone data from a particular mobile operator of unknown market share, and uses a proprietary algorithm to generate their population estimates, precluding any evaluation of primary sources or inherent biases. In both cases, it is impossible to accurately quantify the biases.

Our data show that emigration after disasters has a nonlinear effect on the count of population at-risk and that this emigration is heterogeneous by rurality, affecting the denominators of many key population statistics. As interest in passively-collected data grows and these tools are further refined to overcome current limitations, they can provide a more temporally and spatially nuanced picture of population movements after disasters.

## Methods

### American Community Survey

We obtained yearly population estimates for 2010 to 2018 from the intercensal yearly population estimates provided by the American Community Survey (ACS), a yearly survey conducted by the US Census Bureau. Specifically, we have the vintage 2018 estimates that were published last year.

### Facebook

Facebook is a social media company with over 2 billion monthly active users. In 2017 the Data for Good team within Facebook launched Disaster Maps with the goal of aiding response organizations with information vital to optimal resource allocation in post-disaster settings. These maps are built using privacy-preserving techniques including aggregation and de-identification that protect individual privacy. The Harvard team accessed these data free of charge through Data for Good’s standard license agreement, which allows partners working in humanitarian operations and research to improve their work through use of Disaster Maps.

For this study, we used the original version of Facebook’s *Displacement Maps*, a specific product within Disaster Maps (22), which has since been updated to improve its data sources and methodology. Displacement Maps are generated by first defining a *geographic bounding box* along with an index date defining the disaster event of interest. In this case, the geographic bounding box consisted of the entire island of Puerto Rico and the index date was September 20th, 2017, the day that Hurricane Maria made landfall on the island. In these data, Puerto Rico is divided into 188 non-overlapping *geographical tags (geotags)* and users are assigned a *home location* defined as the geotags where they had the most interactions with the Facebook platform through a browser during the five weeks prior to the index date. In the original Displacement Maps methodology, the location of these interactions were determined from associated internet protocol (IP) addresses; the new methodology instead utilizes location based data from cell phones. Displacement Maps generate a closed population consisting of people using Facebook satisfying the following two conditions: 1) they registered an interaction with Facebook services from in the geographic bounding box during the five weeks preceding the index date, and 2) they were present in their home location during the week before the disaster (23).

They then followed this cohort through July of 2018 and calculated the most commonly occurring geotag each week, aggregating total numbers of cohort members per geotag. Geotags with fewer than 100 people at baseline were excluded to protect privacy. Then for each of the 49 weeks after the index date, the dataset includes the number of crisis-affected people in their home location, the number of new users, and the number of unknown users for each geotag. Cohort members were defined as having an unknown location if they did not register an interaction during the week for which data was aggregated. For our results we assumed these people with unknown locations were in their home location. We then combined geotag counts into Puerto Rico’s 78 municipalities. Due to low counts, the municipality of Las María had no data. Below we describe how we imputed other missing data.

### Teralytics

Teralytics is a tech-company that works with governments and private clients to assess human movement by partnering with mobile network operators. Specifically, the partnership allows the company to access and analyze the data that cell towers receive from mobile devices. From them we obtained island-wide daily population proportion estimates relative to an undisclosed baseline from May 31, 2017 to April 30, 2018 on the basis of all subscribers of a major, undisclosed, telecom company that created events in Puerto Rico. Events were defined as signal exchanges between a cell phone and the nearest cell phone tower. These signal exchanges occurred when a phone call was made, a text message was sent, or any activity that would require a cell phone to connect to a cell tower. The data was filtered by the provider and only subscribers with activity all over the Teralytics analysis period were considered (May 31, 2017 to April 30, 2018). Activity was defined as an event on at least ten days per month for all twelve months. Due to the unreliability of data, proportion estimates for four weeks after hurricane Maria were not provided. For every day, distinct number of subscribers with extrapolated events in Puerto Rico are computed by considering events generated from different mobile devices. Extrapolated events meaning that for any particular daya subscriber is assumed to be on the island if at least one event occurred during that same day on the island and no event occurred outside of Puerto Rico (within the mainland USA).

### Aviation Records

Finally, through the Puerto Rico Institute of Statistics (PRIS) we obtained Airline Passenger Traffic (APT) data from the US Bureau of Transportation Statistics (BTS). The data is composed of monthly counts of passengers that arrived and left the island per month from January 2010 to February 2018. The per-month difference between these two numbers will be referred to as net migration. We added the monthly net migrations to the vintage 2017 population estimates corresponding to the same date and interpolated using a linear model between these data points. This resulted in daily population estimates from July 2010 to July 2018 that accounts for flight passenger movement.

See Supplemental Table 1 for a side-by-side comparison of all the data sources.

### Estimating Population Size

We estimated island-wide population sizes using each of the four data sources. For the ACS data we simply interpolated the points for each year (Supplemental Figure 3). For the other three sources we defined population size estimate *N*_*t*_ for time *t* using the following:

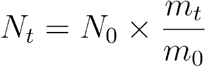

where *N*_0_ = 3,325,001, the ACS population estimate for July 1, 2017, *m*_*t*_ is a source-specific measurement for time *t*, and *m*_0_ is a source-specific baseline. For the APT data baseline was defined as July 1, 2017, *m*_*t*_ correspond to the sum of *N*_0_ and the cumulative net passenger movement for month *t*, and *m*_0_ is the sum of *N*_0_ and the cumulative net passenger movement at baseline. The formula above therefore is simply the cumulative sum, starting on July 1, 2017, of passengers arriving and passengers leaving:

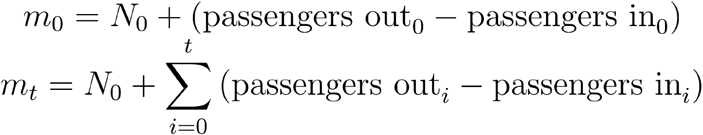

where *i* represents the *i*^*th*^ month after baseline and *Passenger out*_*i*_ and *Passenger in*_*i*_ are the passengers leaving and entering Puerto Rico in month *i*, respectively. For the cell phone data the baseline was also defined as July 1, 2017, *m*_*t*_ and *m*_0_ represent the proportions provided by Teralytics for time *t* and the proportion corresponding to baseline, respectively. Finally, for the Facebook data baseline was defined as August 21, 2017, which corresponds to the first observation in the dataset. Here *m*_*t*_ corresponds to the Facebook population at time *t* and *m*_0_ represents the Facebook population at baseline.

For the municipality level data, we aggregated the city specific data from Displacement Maps by municipality in Puerto Rico while maintaining the temporal resolution at weeks. For all analyses we assumed that cohort members who were defined as *unknown* for the week, were present in their home town locations and were either unable to, or chose not to interact with Facebook services using a browser in that time period.

To evaluate the utility of Displacement Maps, a tool to target municipalities for resource allocation, we evaluated the proportion of the population with unknown locations every week compared to baseline. Our primary assumption here is that in the immediate aftermath of a disaster, any discontinuation of interaction with Facebook services defined at baseline would primarily be caused by loss of access to infrastructure, death or other factors related to the event. Therefore, a higher proportion of municipality specific cohort members whose location was unknown would be a proxy for impacts of the disaster in that region.

### Imputation

As stated before, for privacy-preserving reasons, geotags are excluded from the data if they had fewer than 100 Facebook cohort members. This implies that we did not know the exact total number of users for those geotags. We therefore applied an imputation approach along with a sensitivity analysis to provide intervals of possible values. First, we computed the distribution of the maximum number of consecutive *new*-missing values for each geotag. Thus, the *j*^*th*^ geotag had a corresponding value, *c*_*j*_^(*new*)^, that represents to the maximum number of consecutive weeks where the *new* variable was missing. Second, we denote a threshold **τ** that corresponds to the maximum number of consecutive weeks we are “confident” to accept and a default imputation value **δ** to be used later. Then we split the data into the geotags that comply with the threshold and those that did not (i.e., if *c*_*j*_^(*new*)^ ≤**τ** then geotag *j* complies with the threshold). For the geotags that complied with the threshold, we imputed the missing values using a linear interpolation of the observed values. For the geotags that did not comply with the threshold, we imputed the missing values with **δ**. Finally, we computed the distribution of the maximum number of consecutive *home*-missing values for each tag, in which the *j*^*th*^ geotag had a corresponding value, *c*_*j*_^(*home*)^, and excluded all geotags where the parameter was greater than **τ**. For our results we used **τ**=8 and **δ**=99 (Supplemental Figure 5). Note that **δ**=99 is the maximum number that the missing values can take. We conducted a sensitivity analysis were **δ**=0, the minimum number that the missing values can take (Supplemental Figure 6). Our sensitivity analysis shows that the results do not change much.

## Data Availability

A Github repository with some of the data used in this project will be publish in the future.

## Acknowledgements

We would like to thank Teralytics for providing the mobile phone data used in this project. Specifically, thanks to Canay Deniz, Lara Montini, and Andrea Samdahl for the discussions.

We would also like to thank Facebook for providing the displacement data used in the project. Specifically we would like to thank Shankar Iyer, Laura McGorman, Paige Maas and Facebook’s Data for Good team for lengthy discussions of their algorithm and possible biases.

## Supplemental Material

**Supplemental Figure 1:**
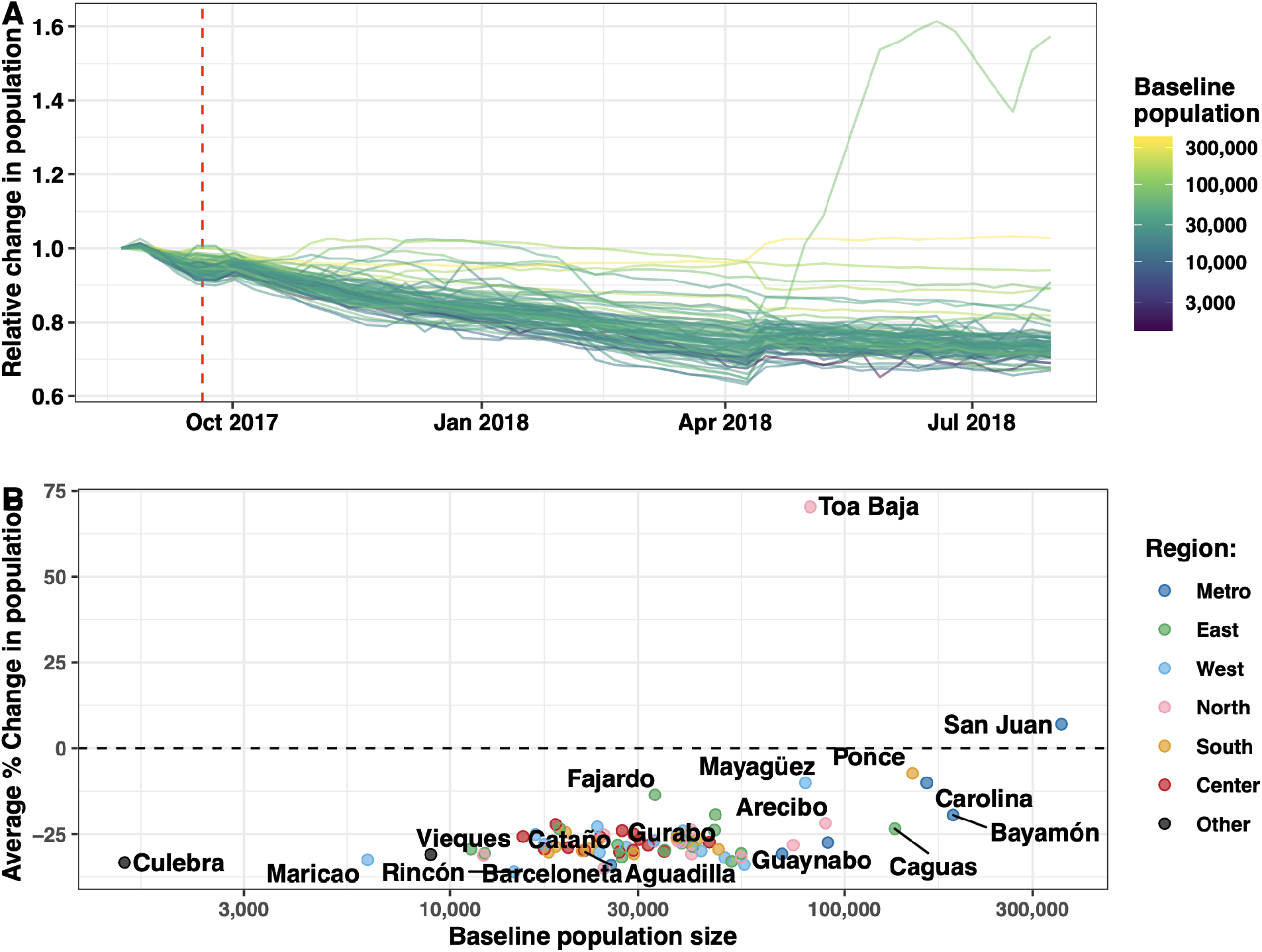
Small towns lost a larger portion of their population compared to large ones. This figure is the same as Figure 2 but includes the municipality of Toa Baja (A) Relative change in the population of Facebook cohort member per municipality. Each curve corresponds to a different municipality, Population size in July 2017 is denoted by color. (B) Average percent change in population at the end of the study period, relative to baseline, compared to the ACS population size of municipality at baseline. The geographical regions are presented in Supplemental Figure 4. We fitted a linear model to each population curve and computed the average percent change using the first and last fitted values. The vertical lines correspond to the day when Hurricane Maria made landfall in Puerto Rico.

**Supplemental Figure 2:**
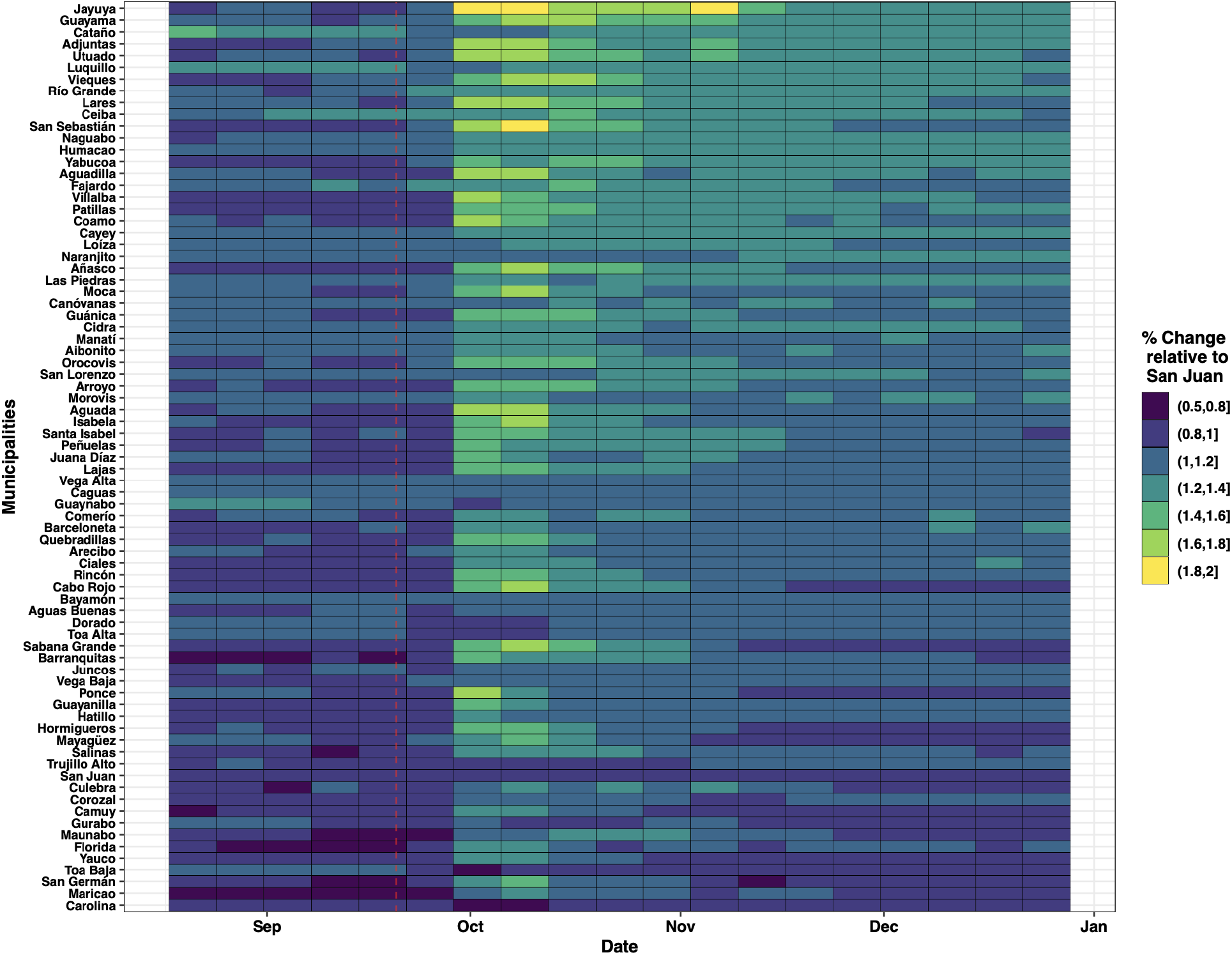
Proportion of Facebook cohort members in Puerto Rico whose location was unknown by municipality compared to the same proportion in San Juan sorted by population at baseline. The red line corresponds to Hurricane Maria’s landfall.

**Supplemental Figure 3:**
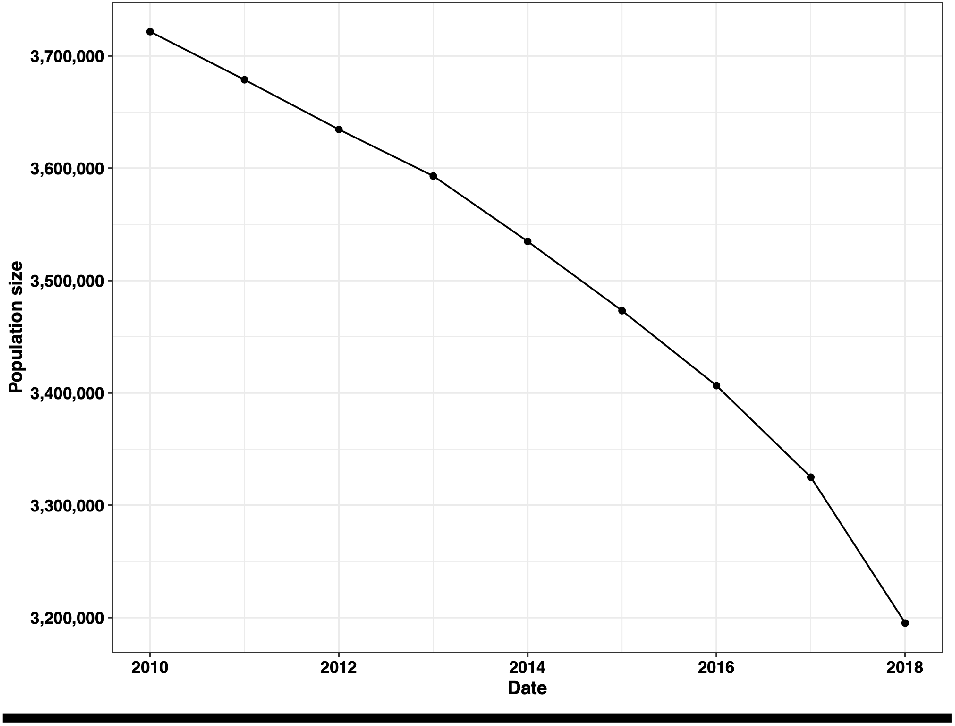
US Census population estimates for Puerto Rico from 2010 to 2018

**Supplemental Figure 4:**
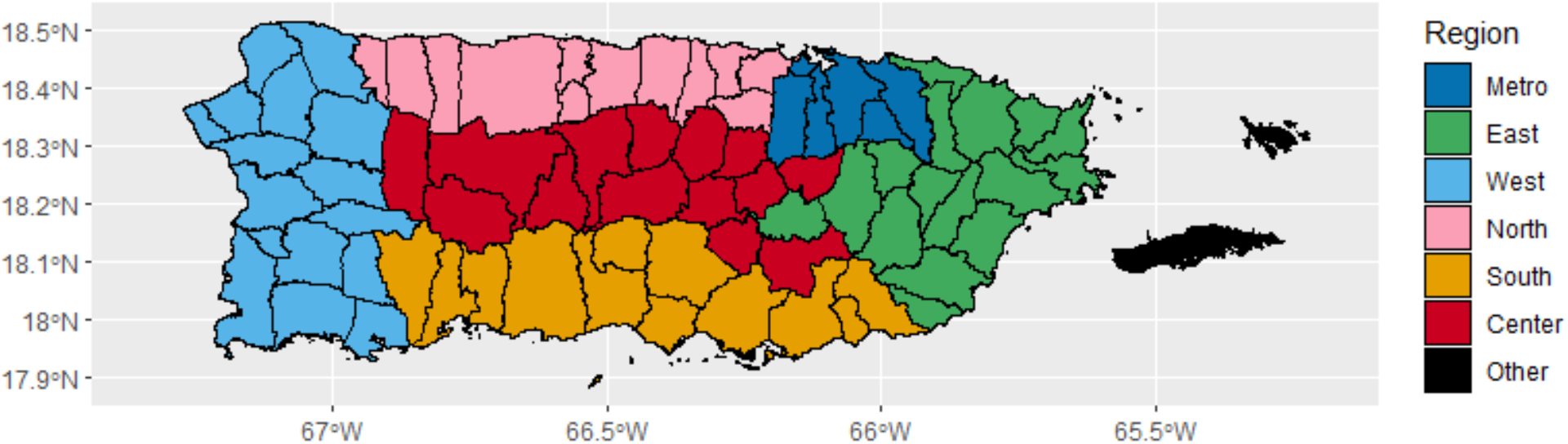
Regions of Puerto Rico

**Supplemental Figure 5:**
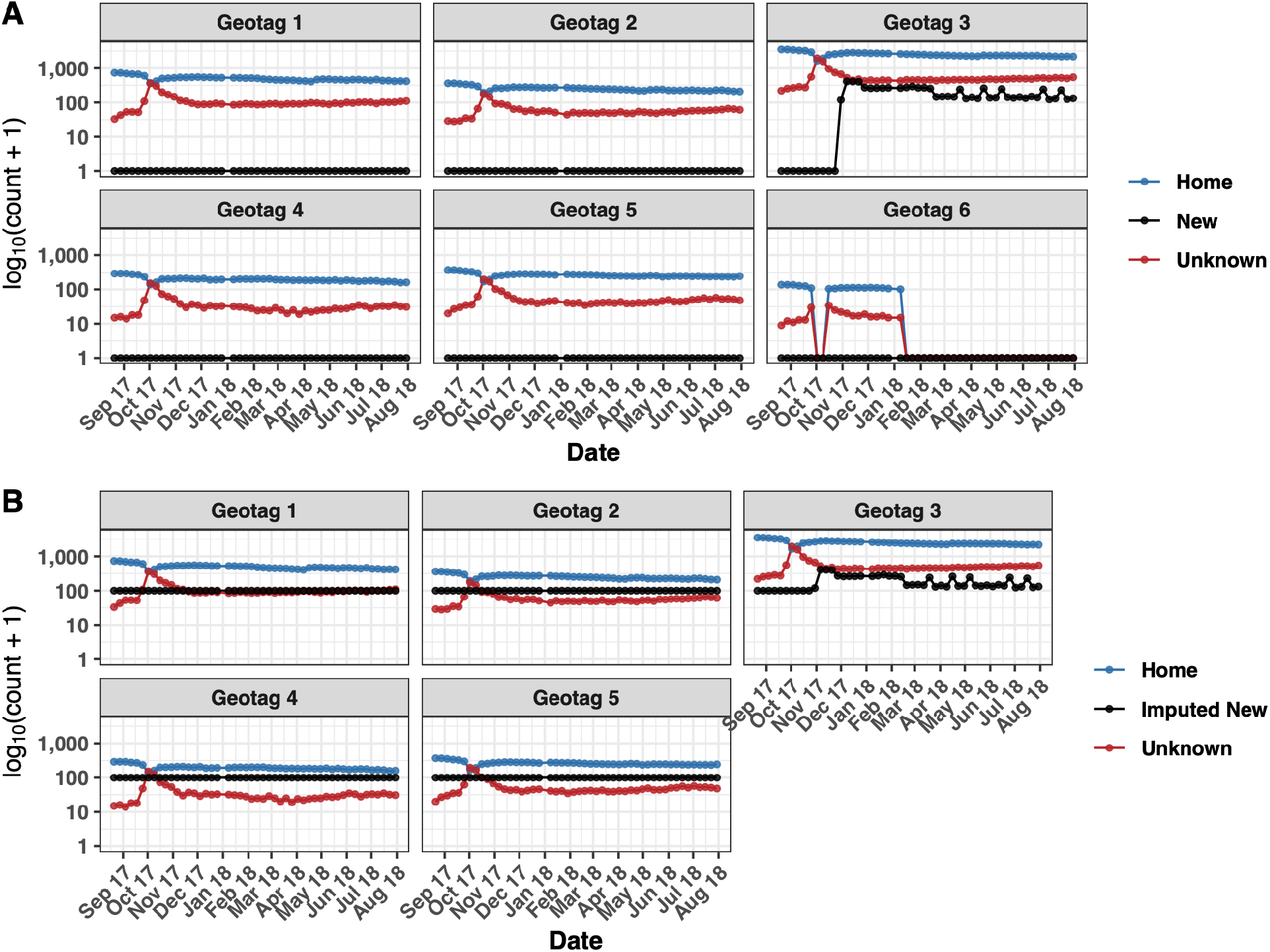
Results of our imputation technique with **τ** = 8 and **δ** = 99 for the municipality of San German. (A) Geotags for the municipality of San German showing data for individuals remaining in their home location, new individuals arriving at locations in San German, and individuals from locations in San German whose current location is unknown at each time point. (B) Results of our imputation technique applied to the municipality of San German. Note that **Geotag 6** is not included since it does not comply with the threshold **τ** and the imputation of **New** with the default imputation value **δ**.

**Supplemental Figure 6:**
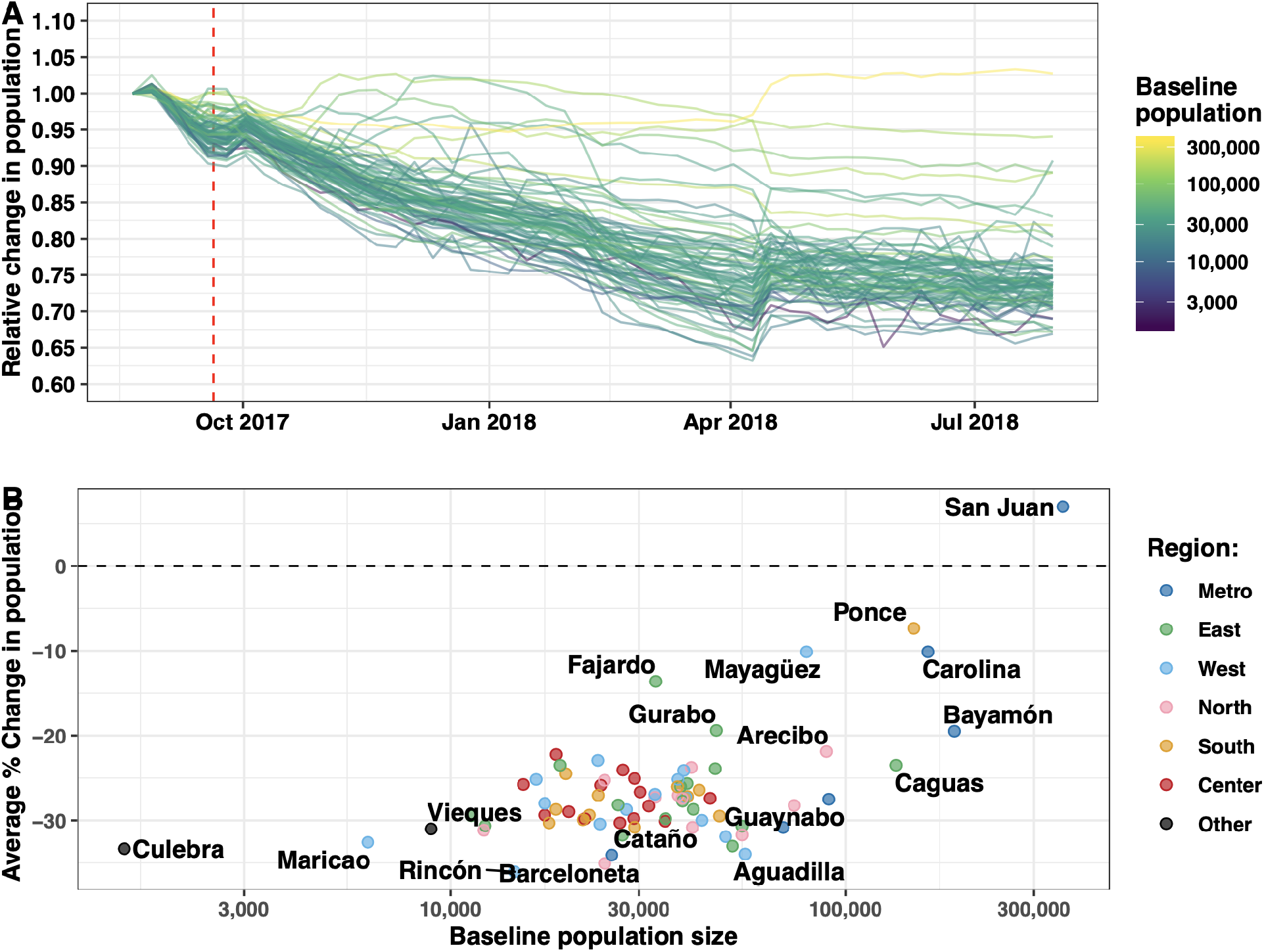
Results of our imputation technique with **τ** = 8 and **δ** = 0. Small towns lost a larger portion of their population compared to large ones. For a reference, compare this to Figure 2. (A) Relative change in the population of Facebook cohort member per municipality. Each curve corresponds to a different municipality, Population size in July 2017 is denoted by color. (B) Average percent change in population at the end of the study period, relative to baseline, compared to the ACS population size of municipality at baseline. The geographical regions are presented in Supplemental Figure 4. We fitted a linear model to each population curve and computed the average percent change using the first and last fitted values. The vertical lines correspond to the day when Hurricane Maria made landfall in Puerto Rico.

**Supplemental Table 1:** Comparison of all used data sources

| **Table 1: Comparison of Data Sources (Continued)** |  |  |  |  |
| --- | --- | --- | --- | --- |
| **Data Source** | **Granularity** | **Utility** | **Foreseen Problems** | **Anticipated Direction of Bias** |
| Aviation Records | Temporal: Monthly<br>Geographic: Island-wide | Useful to capture population fluctuations in regions without significant migration outside of aviation. | - Captures total number of passengers, not specifically Puerto Ricans.<br>- Unable to identify if same individuals leave and come back. | Unknown. May potentially overestimate the true population size as the passengers may include those not from Puerto Rico and those who were not on the island before Hurricane Maria. |
| Census (ACS) | Temporal: Yearly<br>Geographic: Municipality | Most valid estimate of estimated population size between census years. | - Is often published months after a disaster period.<br>- Provides a single estimate with a confidence interval for the entire year.<br>- Is updated year on year as more information becomes available. | Unknown. In the immediate aftermath of a disaster the monthly samples may more heavily weight data available from pre-disaster points of time providing an overestimate of the true population size. |
| Facebook | Temporal:<br>Weekly<br>Geographic:<br>Cities as defined by Facebook | Available immediately with a transparent counting algorithm and representative of the population which experienced the disaster. | - Only captures individuals who use Facebook through a browser which may be heterogeneous by age groups and socio-economic status.<br>- Cohort will generally decrease as membership is defined by presence in Puerto Rico in the five weeks before Hurricane Maria.<br>- Counting algorithm will exclude towns where there are fewer than 100 individuals disproportionately affecting rural areas.<br>- Dependent on access to electricity and internet. | Will likely underestimate the true population size as the closed cohort will only register out-migration and deaths while in-migration and new users are ignored. |
| Call Detail Records | Temporal:<br>Daily<br>Geographic:<br>Island-wide | Not dependent on surveys or users of an application. Assuming that usage of a specific telecommunications provider is random, provides a large representative sample of island population size. | - Counting algorithm is not clear and proprietary.<br>- Dependent on access to electricity and cell phone coverage. | Unknown. Due to the lack of information on the telecommunications operator providing the sample of Puerto Ricans or the extrapolation algorithm, it is difficult to evaluate any potential bias. |

**Supplemental Table 2:** Top 15 municipalities in Puerto Rico with greatest proportion of Facebook cohort members whose location was unknown in the week following Hurricane Maria

| Municipio | Date | Proportion Unknown | San Juan proportion unknown for week i | Ratio of Proportion Unknown |
| --- | --- | --- | --- | --- |
| Jayuya | 10/2/2017 | 0.660598 | 0.365703 | 1.80638 |
| San Sebastián | 10/2/2017 | 0.654775 | 0.365703 | 1.790456 |
| Utuado | 10/2/2017 | 0.63819 | 0.365703 | 1.745106 |
| Adjuntas | 10/2/2017 | 0.631111 | 0.365703 | 1.725749 |
| Lares | 10/2/2017 | 0.622645 | 0.365703 | 1.7026 |
| Villalba | 10/2/2017 | 0.60396 | 0.365703 | 1.651506 |
| Ponce | 10/2/2017 | 0.603934 | 0.365703 | 1.651434 |
| Aguadilla | 10/2/2017 | 0.596493 | 0.365703 | 1.631087 |
| Aguada | 10/2/2017 | 0.593456 | 0.365703 | 1.622781 |
| Coamo | 10/2/2017 | 0.592866 | 0.365703 | 1.62117 |
| Isabela | 10/2/2017 | 0.583985 | 0.365703 | 1.596885 |
| Peñuelas | 10/2/2017 | 0.582012 | 0.365703 | 1.59149 |
| Juana Díaz | 10/2/2017 | 0.580964 | 0.365703 | 1.588623 |
| Añasco | 10/2/2017 | 0.579461 | 0.365703 | 1.584514 |
| Sabana Grande | 10/2/2017 | 0.567899 | 0.365703 | 1.552899 |

## References

1. African Development Bank Group, L. Mbaye, Climate change, natural disasters, and migration. IZA World Labor (2017) https:/doi.org/10.15185/izawol. 346 (June 11, 2019).

2. D. Thomaz, Post-disaster Haitian migration. States Fragility, 2.

3. K. J. Curtis, E. Fussell, J. DeWaard, Recovery Migration After Hurricanes Katrina and Rita: Spatial Concentration and Intensification in the Migration System. Demography 52, 1269–1293 (2015).

4. The National Institute for Occupational Safety and Health (NIOSH), Methods: Mortality. CDC Append. (June 17, 2019).

5. F. Checchi, L. Roberts, Interpreting and using mortality data in humanitarian emergencies. A primer for non-epidemiologists. Humanit. Pract. Netw. 44 (2005).

6. United States Census Bureau, “Methodology for the Intercensal Population and Housing Unit Estimates: 2000 to 2010” (June 11, 2019).

7. L. Bengtsson, X. Lu, A. Thorson, R. Garfield, J. von Schreeb, Improved response to disasters and outbreaks by tracking population movements with mobile phone network data: A post-earthquake geospatial study in haiti. PLoS Med. 8 (2011).

8. L. Bengtsson, et al., Using Mobile Phone Data to Predict the Spatial Spread of Cholera. Sci. Rep. 5 (2015).

9. NOAA, “Tropical Cyclone Report Hurricane Maria (AL152017)” (May 15, 2019).

10. C. D. Zorrilla, The View from Puerto Rico — Hurricane Maria and Its Aftermath. https://doi.org/10.1056/NEJMp1713196 (2017) https:/doi.org/10.1056/NEJMp1713196 (May 15, 2019).

11. C. Arnold, Death, statistics and a disaster zone: the struggle to count the dead after Hurricane Maria. Nature 566, 22 (2019).

12. N. Kishore, et al., Mortality in Puerto Rico after Hurricane Maria. N. Engl. J. Med. (2018) https:/doi.org/10.1056/NEJMsa1803972 (May 15, 2019).

13. J. Sandberg, et al., All over the place? Differences in and consistency of excess mortality estimates in Puerto Rico after hurricane Maria. Epidemiology Publish Ahead of Print (2019).

14. R. Cruz-Cano, E. L. Mead, Excess Deaths After Hurricane Maria in Puerto Rico. JAMA 321, 1005–1005 (2019).

15. A. R. Santos-Lozada, J. T. Howard, Use of Death Counts From Vital Statistics to Calculate Excess Deaths in Puerto Rico Following Hurricane Maria. JAMA 320, 1491 (2018).

16. R. Rivera, W. Rolke, Modeling excess deaths after a natural disaster with application to Hurricane Maria. Stat. Med. 38, 4545–4554 (2019).

17. United States Census Bureau, More Puerto Ricans Move to Mainland United States, Poverty Declines. U. S. Census Bur. (December 20, 2019).

18. J. Schachter, A. Bruce, The Impact of Hurricane Maria: (2019).

19. R. Á. Claudio, 10 municipios de Puerto Rico declarados zona de desastre. Metro (June 17, 2019).

20. M. Duggan, J. Brenner, The Demographics of Social Media Users — 2012. 14.

21. NapoleonCat, “Facebook users in Puerto Rico - October 2018” (Facebook API) (December 19, 2019).

22. Facebook Data for Good, Using Data to Help Communities Recover and Rebuild | Facebook Newsroom (May 14, 2019).

23. Facebook Data for Good, Facebook Disaster Maps: Methodology. Facebook Res. (2017) (May 14, 2019).

